# Knowledge Driven Phenotyping

**DOI:** 10.1101/19013748

**Authors:** Honghan Wu, Minhong Wang, Qianyi Zeng, Wenjun Chen, Jeff Z. Pan, Cathie Sudlow, Dave Robertson

**Affiliations:** The University of Edinburgh; The University of Aberdeen; Health Data Research, University of Edinburgh, United Kingdom

**Keywords:** health data, phenotype computation, data integration, ontology

## Abstract

Extracting patient phenotypes from routinely collected health data (such as Electronic Health Records) requires translating clinically-sound phenotype definitions into queries/computations executable on the underlying data sources by clinical researchers. This requires significant knowledge and skills to deal with heterogeneous and often *imperfect* data. Translations are time-consuming, error-prone and, most importantly, hard to share and reproduce across different settings. This paper proposes a knowledge driven framework that (1) decouples the specification of phenotype semantics from underlying data sources; (2) can automatically populate and conduct phenotype computations on heterogeneous data spaces. We report preliminary results of deploying this framework on five Scottish health datasets.

## 1. Introduction

Big data analytics in healthcare has great potential to reveal deep insights from health data, which would extend our knowledge boundary in medicine and improve quality of health service [1]. However, it is very challenging to make sense of distributed and heterogeneous health data. The current reality is that most data is stored in different local communities, which means they are maintained locally and stored in inconsistent formats and languages. A key technical challenge haunting almost all data-driven clinical studies is to extract or compute accurate patients’ phenotypes (traits of symptoms, diseases, medications or biochemistry test results) from such a fragmented data space.

Figure 1 (*Current Practice section* on the left-hand side) illustrates a typical procedure of computing phenotypes from heterogeneous data sources. The first step, and scientifically the most important, is to specify what constitutes a phenotype using clinical knowledge/terminologies that the clinical research community are familiar with, e.g. using languages in laboratory, medicine, oncology and genetics etc. These specified phenotypes must be computed (extracted or inferred) from the underlying health data. Their computation (steps 2 & 3) requires (each time) significant human effort to understand database details, good data science skills to do the querying and data manipulating, and caution & patience to deal with data incompleteness/inconsistencies. The translations from phenotype specifications into computations are hard to verify and debug, which poses threats to the understandability and credibility of data-driven studies. In addition, due to the sensitive nature of health data, such translations cannot be made public, which significantly impedes the reusability and reproducibility of clinical researches. In this paper, we propose a knowledge driven framework to tackle these challenges.

**Figure 1.**
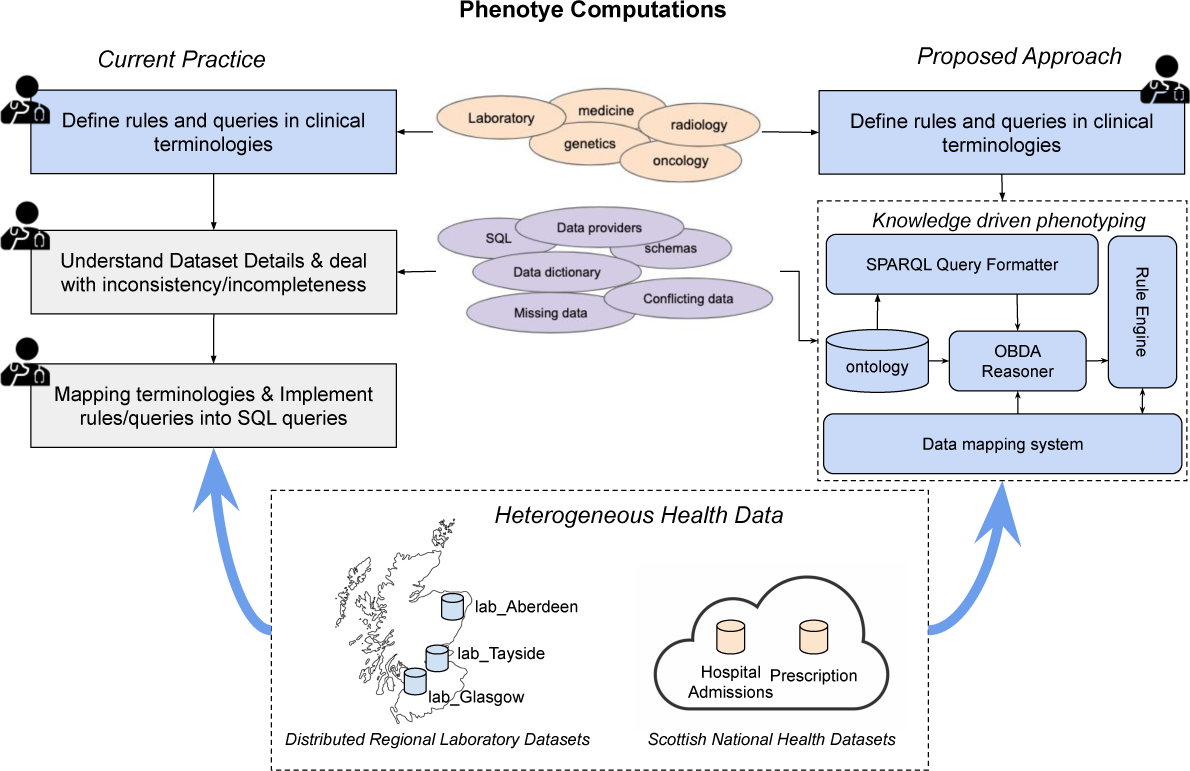
System architecture. A knowledge driven phenotyping framework that frees clinical researchers from time-consuming and error-prone translations from clinical rules to database-level queries.

## 2. Method

The main aim of this study is to realise a clinical data science framework that makes the underlying data sources *transparent* to phenotype computations. Researchers only need to specify the “meanings” of their phenotypes using their familiar terminologies and the actual computations are automatically populated for and executed on data sources.

For example, to retrieve a cohort of patients with Type 2 Diabetes in Scotland, such a framework would only require researchers to provide an assertion like *Type*2 *Diabetes Patient*(?*x*) without them knowing that the national *Hospital Admissions* is the underlying database, which is a Microsoft SQL Server managed by eDRIS team (https://www.isdscotland.org/Products-and-Services/eDRIS/) (therefore, they have the data dictionary) and uses both ICD 9 and 10 for diagnosis coding due to legacy data. To realise that, the key is to decouple the formalisation of phenotype semantics and the management/technical details of underlying data sources. We propose an architecture (see Figure 1) to implement such a decoupling, which is roughly composed of two aspects: phenotype formalisation framework (for specifying phenotype semantics) and ontology/rule based data access (for automated phenotype computing on data sources).

### 2.1. Phenotype Formalisation Framework

Given a phenotype such as *Type*2_*Diabetes_Patient*, the computer has to understand its *semantics* (i.e., computer understandable meanings) so that the right computations and queries can be populated and executed on the underlying data sources. The formalisation framework has three components as follows.

- A database independent phenotype formalisation using Semantic Web knowledge representation technologies. Specifically, we use DL Lite [2] ontologies and Semantic Web Rule Language (SWRL) [3] queries to define phenotype semantics.
- A core phenotype ontology serving as the base vocabulary linking to standard clinical terminologies available at BioPortal [4] (e.g., SNOMED CT, ICD10).
- A query formatter that generates ontology queries (SPARQL [5]) from an user interface. The formatter can automatically translate phenotype definitions between standard terminologies (e.g., READ to SNOMED CT) via UMLS [6].

We use *Type 2 Diabetes* as a running example to illustrate phenotype specifications in this framework.

*Phenotype Definition using Ontology* The following equation defines a DL-Lite axiom for computing the phenotype of Type 2 Diabetes, meaning anyone who had a diagnosis of ICD10 code E11 is a Type 2 Diabete Patient.

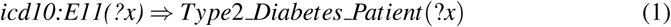

*Standard Terminology Inclusion and Subsumption Inference* The prefix *icd10:* indicates that *E11* is a concept of ICD10 ontology, which is then incorporated in the phenotype computing. This automatically includes all the *semantics* defined in ICD10, such as *icd10:E11.2 ⊑ icd10:E11*. Based on DL-Lite logic, such a class subsumption relation will be automatically combined with Axiom (1) to infer that *icd10:E11.2 Type*2 *Diabetes Patient*. Similarly, instances of all other sub-classes of *E11* are inferred as instances of *Type*2 *Diabetes Patient*.

*Rule for Dealing with Incompleteness* If a researcher worries the diagnosis data might miss some Type 2 Diabetics, she might think of certain drug uses which could infer the disease phenotype with acceptable reliability. Rule (2) infers that a patient prescribed *Sulphonylurea SR* is potentially a Type 2 Diabetes patient.

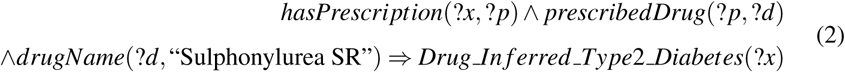

*Rule for Dealing with Inconsistency* Erroneous and inconsistent facts are almost inevitable when using clinical data for research. For example, clinically, it is very unlikely a person is both Type 1 and Type 2 Diabetic. The rule and axiom in (3) define a class of *DiabetesCon f lictPatient* as a sub-class of *PotentialCodingErrorPatient* in the core ontology, which detects potential coding errors in health data.

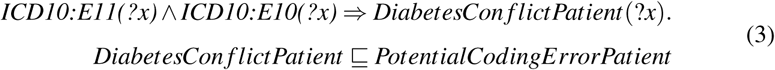

### 2.2. Ontology Based & Rule Driven Data Access

To *automatically* compute the above formalised phenotypes on actual data, we adopt ontology based data access (OBDA) techniques [7]. The OBDA system in our framework (on the right of Figure 1) is composed of three components: an **ontology** for specifying user queries, a **data mapping system** for translating ontology predicates into relational database schema/constructs and an **OBDA reasoner** conducting the translation and optimisation, for which we use an open source OBDA reasoner - OnTop [8].

Phenotype computations are heavily rule driven for two reasons: (1) most phenotyping semantic constraints are hard to represent using OWL Lite constructs (e.g., the rules described previously); (2) the semantics of most phenotypes are not fixed - they either change with research focuses or different researchers might have different opinions about certain rules related to a phenotype. Therefore, comparing to ontological formalisation, phenotype semantics needs more customisable/flexible specification approaches like rule languages. However, the current official release of OnTop does not support SWRL rules.

For this reason, in our framework, we minimise the core ontology. A core mapping from this ontology to the underlying data sources is manually created to initiate the data mapping system. To support rule-driven phenotype specification, a **rule engine** is implemented. The engine can automatically convert SWRL rules into new data mappings by utilising the core mapping and the ontology semantics. The populated mappings are then merged with the core mapping in the rule engine, which will be loaded in the OBDA component on the fly to do phenotype computations.

## 3. Deployment and Evaluation

This study is supported by Health Data Research UK (https://www.hdruk.ac.uk/projects/graph-based-data-federation-for-healthcare-data-science/) as an exemplar to create a federation of distributed health data in Scotland. The above described framework has been deployed on 5 synthetic data sets (as shown at the bottom of Figure 1) generated using BadMedicine (https://github.com/HicServices/BadMedicine), which represents data/schema characteristics learnt from real data.

Due to space limitations, we put the full benchmark and evaluation details on a Github page: https://github.com/Honghan/KGPhenotyping/tree/master/evaluation. In Table 1, we compares the ontology queries (SPARQL) and SQL queries for three exemplar phenotype definitions (in simplified forms; actual queries are at https://bit.ly/2oZK5rK).

**Table 1.**
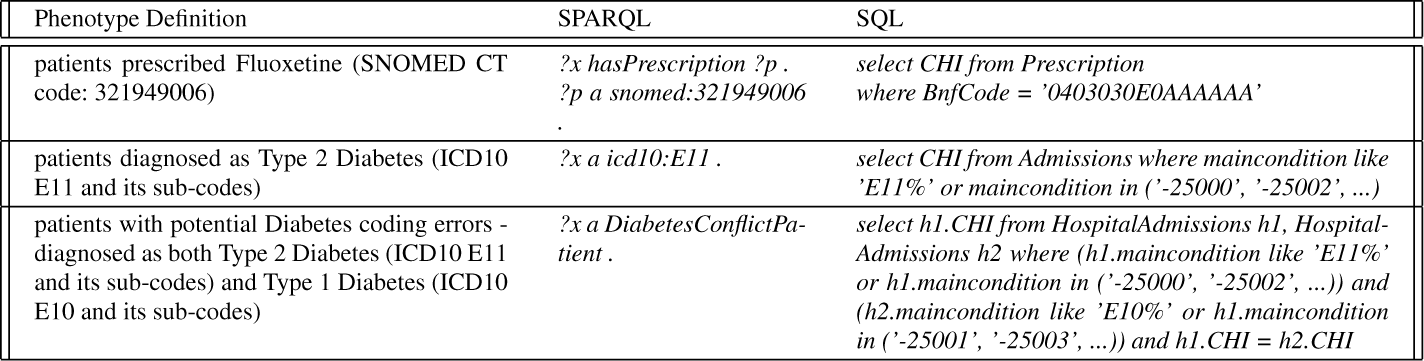
SPARQL vs. SQL queries for the three phenotype definitions on 5 Scottish Health Datasets

1. *(Terminology Mapping)* The first phenotype is defined using SNOMED CT (321949006) to specify a prescription. Scottish Prescription data is using Bnf-Code for drug identifiers. Our framework can take the SNOMED CT code as the input and translate it into BnfCode automatically, while in Current Practice the user has to understand that BnfCode is used in the data and need to get equivalent BnfCode (0403030E0AAAAAA) for the given code.
2. *(Inference)* The second phenotype is to get patients with Type 2 Diabetes defined as ICD10 E11 and its sub-codes. However, in Scottish Hospital Admissions dataset, both ICD10 and ICD9 are used. Therefore, using SQL, it requires us to give an exhaustive list of codes (20 codes). In our framework, the ICD9/10 hierarchies and their mapping are automatically utilised for doing inferences. Therefore, only a simple *icd10:E11* predicate will be sufficient for the computation.
3. *(Inconsistency Checking)* The third phenotype is to detect potential Diabetes coding errors in the data (Type 1 and 2 are exclusive). With Rule 3, our framework only needs one predicate *DiabetesCon f lictPatient*, while SQL will need around 40 codes and a inner join operation on the Hospital Admission table.

## 4. Conclusion

To overcome *obscure* phenotype computation, which makes experiments difficult to understand and reproduce, we developed a new framework to allow clinical researchers to formalise phenotype semantics independently to the data and, more importantly, in a computer understandable way so that its computation can be automated on the underlying data sources. We implemented a knowledge-driven (based on ontologies and rule languages) approach to define an interlingua in which practitioners can represent the phenotype semantics they want to use and automatically translates this to computations as database queries on participating data sources.

## Data Availability

The research is conducted on national and regional health data in Scotland. Given the privacy/sensitivity of the data, we cannot share the actual health data. However, synthetic data can be generated using BadMedicine (https://github.com/HicServices/ BadMedicine). It uses the actual data schema and represents the distributions learned from the real data.

## Notes

### Competing Interest Statement

The authors have declared no competing interest.

### Funding Statement

This project is funded by MRC/ISCF HDRUK(Grand no: MC_PC_18029) and MRC/HDRUK (Grand no: MR/S004149/1).

